# Effectiveness of mHEALTH Application at Primary Health Care to Improve Maternal and New-born Health Services in Rural Ethiopia: Comparative study

**DOI:** 10.1101/2022.04.02.22272628

**Authors:** Aragie Kassa, Mokgadi C. Matlakala

**Affiliations:** Siba Consulting, Addis Ababa, Ethiopia; Department of Health Studies, University of South Africa, Pretoria, South Africa

## Abstract

Ethiopia has recently implemented mHealth technology on a limited scale to help increase the uptake of health services, including intervention for maternal and new-born health service utilisation. In this study, the effectiveness of the mHealth intervention was assessed by measuring the level of maternal health service utilization in 4 Health Centers in Ethiopia.

The study was comparative by design employing comparison of maternal and newborn health service utilization before and after initiation of mHealth implementation. Follow-up data of 800 clients were randomly selected and included in the study, to determine the magnitude of maternal and new-born health service utilization. Data analysis included comparison of pre-mHealth (baseline) with mHealth follow-up data, using independent t-test to compare magnitude of maternal and new-born health service utilization.

The mean of antenatal care follow-up during their recent pregnancy was 2.21(SD±1.02) and 3.43(SD±0.88) for baseline and intervention, respectively. Antenatal visit of four or more was reached for 55(13.8%) of the baseline and 256(64%) of pregnant women in the mHealth intervention group. Pregnant women’s timeliness to start ANC follow-up at baseline and intervention groups was 44.5% and 77.3%, respectively. Institutional delivery at baseline and intervention groups were 35.0% and was 71.2%, respectively. Of women who gave birth, 23.8% at baseline received first postnatal care within 6 hours, 11.3% within 6 days, and 6.8% within 6 weeks. In the intervention group, 84% delivered women received first postpartum within 6 hours after delivery, 70.8% after 6 hours, and 46% made their third postpartum visit within 6 weeks after delivery. Penta-3 vaccination coverage at baseline and mHealth intervention groups was 61.5% and 70.4%, respectively.

The study result suggest that the introduction of a low-cost mHealth technologies contributed to the observed improvement of maternal and new-born health service utilization. This intervention shows promise for scale up as well as to be applied to other health interventions beyond maternal and newborn health services.

## INTRODUCTION

World Health Organization (WHO) disclosed death of 810 women per day in 2017 due to complications to pregnancy and childbirth. Attributable to this, close to 295,000 women died during pregnancy and childbirth in 2017. Most of these deaths (94%) occurred in low-resource constrained countries, and majority could have been prevented. The majority (254, 000) of the deaths occurred in Sub-Saharan Africa and accounted for roughly 77.2% of maternal deaths [1].

The vast majority maternal deaths in some regions of the globe reveals disparities in access to health services and showed the gap between rich and poor. The maternal mortality ratio (MMR) in in resource constrained countries and developed nations in 2017 was 462 per 100, 000 live births and 11 per 100 000 live births, respectively [2].

Since 2000 considerable progress has been made in decreasing child mortalities. The decline of global under-5 mortality rate by 49% in 2000 from 77 deaths per 1000 live births to 39 in 2017. This is comparable to 1 in 14 children dying before reaching age 5 in 2017, compared with 1 in 13 dying before age 5 in 2000[1].

As indicated by WHO, global under five mortality rates declined from 31 per 1000 live births in 2000 to 18 in 2017 in the first month of life, which is a decrease by 41%.The observed fell of death is smaller compared with the 54% reduction in mortality for children aged 1–59 months. Under-5 mortality rates are highest in the WHO African Region and in low-income countries, where one child dies out of 14 born [2].

In Ethiopia, the magnitude of maternal mortality and morbidity are among the highest in the world. Maternal Mortality Ratio (MMR) of 412 per 100,000 live births was documented in 2016[3]. Delivery by skilled birth attendants (SBAs) is endorsed to reduce maternal and neonatal mortality. As indicated in Health Sector Transformation plan (HSTP) a set of high impact interventions were being effected, including antenatal care (ANC), skilled birth services and postnatal (PNC) with the intention of reducing maternal mortality to 267 per 100,000 live births [4]. Although the proportion of pregnant women who received ANC services at least once exceeded 90%, continuity of service and quality of care is not optimal as evidenced by low coverage of skilled delivery, tetanus toxoid (TT) vaccine uptake, and screening for syphilis as well as suboptimal uptake of prevention of mother-to-child transmission of HIV (PMTCT) services by pregnant women [4].

The objectives of the study were to compare the degree of maternal and new-born service utilizations before and after start of mHealth implementation, namely, antenatal care follow-up at least four times, institutional delivery, post Natal care (PNC), and penta 3 immunizations. It also include describing maternal and child morbidity before and after mHealth implementation.

## DESIGN OF mHealth IN HABRU DISTRICT, NORTH WOLLO ZONE

The main purpose of the mHealth system in the study setting was to improve communication between the lower and mid-level health workers at health post and Health Center, respectively. It also includes tracking lost for follow-up clients registered for the first service. On the whole, it is maternal and new-born tracking and follow-up system deployed at the Health Centers selected for the study [5].

The theoretical framework of the mHealth project under implementation was articulated on tracking of pregnant women and narrowing communication gap between the Health Centers and health posts related with referral and follow-up [5]. The web based application system deployed at the Health Center track ANC, delivery, PNC, and pentavalent immunization service utilizations. This tracking system has two components, namely, Web application and SMS engine. The web based application permit the midwife at the Health Center to register new pregnant women and follow them for ANC, delivery, PNC, and child immunization services through the graphical user interface. The SMS engine serves as a communication tool between the midwife and the HEW using text message [5].

The web based artefact at the Health Center level track the following functionalities through the SMS system. The system generates four SMS reminders in connection with ANC visit based on calculation of gestational age or fundal height of pregnant women. Visiting pregnant women at Health Center is marked with identification number (ID). Then, four SMS reminders marked with the ID number of the pregnant woman is scheduled to be dispatched to the cell phone of the HEW at gestational age of 26, 32,36, and 39 weeks [5].

The system sends one SMS at 30, 14, 6, and 2 weeks of expected date of delivery (EDD) to HEW to monitor the pregnant women at home based on the EDD. The HEW report to the Health Center the status of the pregnant women using the women’s ID number. Following delivery the status of the mother in the system is changed to ’delivered’ and system calculates the schedule for post-natal care and sends an alert SMS to the HEW at day 1, day 3, and day 7 for the first, second, and third visits, respectively. Three scheduled SMS at 6th, 10th, and 14th weeks after delivery is sent to the HEW as reminder for follow-up of pentavalent vaccination for the new-born [5].

## Methods

### Study setting

The settings for this study were 4 Health Centers implementing mHealth in Habru District, North Wollo Zone of Amhara Regional State. The intervention group consists of pregnant women who used maternity services (ANC, institutional delivery, post-natal care, and penta-3 immunization) after the start of mHealth and the comparison group were pregnant women attended identical services 24 months prior to the start of mHealth in same Health Centers. Clinical records were maintained on all maternal and new-born health clients used services on entry to ANC and this information was transferred on a daily basis to a computer database.

### Study design

The study was before-and-after comparative design and collected longitudinal follow-up obstetric data of women who used ANC, institutional delivery, PNC, and enta-3 immunization from their obstetric record. The obstetric data of women who used services 24 months following initiation of mHealth were captured from mHealth data base in 2018. Similarly, data of 24 months before start of mHealth was captured to determine magnitude of utilization of health services such as ANC, health facility delivery, postnatal care, and penta 3 immunizations.

The MNH records contain client histories such as obstetric, information on current pregnancy, general medical conditions, evaluations on HIV sero-status, and HIV care. The system also archives information on pregnancy follow-up during the time of at least four ANC visits in quantifiable form. Other descriptive data were also captured from both intervention and baseline groups to further explore the comparability of their background characteristics.

Pre- and post-intervention data was collected using a structured data collection checklist. The data was checked for completeness, coded and entered in to CSPro (Census and Survey Data Processing). Following data cleaning all statistical analyses were performed using Stata 14.2. Statistical calculations were made for both descriptive and inferential statistics, and the results presented in the form of graphs and tables.

### Outcome variables

The outcome of interest for all women were based on four for key outcome indictors: the primary outcomes of this study was utilization of ANC 4 and above, health facility based delivery, number and timing of PNC, and utilization of penta-3 immunizations. Number of ANC visits was measured by adding up all ANC follow-ups as recorded in mHealth database and client records. Likewise, number of institutional deliveries and postpartum visits and number of penta-3 immunizations were captured from mHealth data base and registers.

### Explanatory variables

The study collected data on mother’s age, marital status, HIV prevention and care, women’s clinical history. The study participants were categorized by age, and other obstetric and general health conditions (parity, gestational weight, birth weight, preterm, and others) and presence or absence of co-morbidities.

### Sample size

For this study sample size was computed using two proportions sample size calculation with a power of 80% and 5% level of significance. The estimate for P_1_ is coverage for one time ANC (25%) for Amhara Region as obtained from Ethiopian Demographic and Health Survey (EDHS) 2016. The study used 50% estimate for P_2_ as there is no up-to-date ANC data. Considering the above assumptions the calculated unadjusted sample size was 800. Hence, the sample size for pre-interventions was 400; and that of post-intervention was 400. Therefore, 400 client who used maternal and child services 24 months before the start of mHealth were randomly selected and included for the baseline and 400 women who used maternal and child health services after the start of mHealth were also randomly selected form the mHealth database and included for the post-intervention phase. The calculated sample size was proportionally allocated to each Health Centers based on volume of ANC client flow as indicated in Table 1.

**Table 1:** Distribution of sample size to Health Centers included in the study.

| Health Center | Allocated sample size | % distribution |
| --- | --- | --- |
| Mersa | 288 | 36.0 |
| Wurgesa | 197 | 24.6 |
| Hara | 174 | 21.8 |
| Sirinka | 141 | 17.6 |
| **Total** | **800** | **100** |

### Sampling Technique

The researchers used the probability proportional to size (PPS) technique to select participants from each Health Center and employed simple random sampling technique to select client charts from each Health Center. From a complete list of all mothers meeting eligibility criteria, their card number was captured from the ANC logbook and their chart was selected randomly for review. Using the list of ANC identification number in the ANC logbook as a sampling frame, client identification number (ID) was randomly selected via random number generator. Subsequently, client individual record was traced using those generated numbers.

### Eligibility Criteria

The main inclusion criterion for women and health facilities in the study were

- Health Centers implementing mHealth based maternal and new-born health at least for 24 months prior to data collection,
- Pregnant women irrespective of past pregnancy outcome, who used maternal and new-born health services 24 months before and after the start of mHealth in the selected Health Centers became eligible for this study.

### Data management and analysis

Data were cleaned and appended and a new data set was created that contain the baseline and end line data, with a total of 800 participants. Targeted indicators were presented with 95% confidence interval. Calculated values of same indicators from before and after interventions were compared using independent t-tests. Binary logistic regression models were performed to obtain adjusted odds ratio (OR) for the outcome variables using demographic and clinical variables to estimate predictors of both maternal and new-born health service utilizations, morbidities and pregnancy outcomes. Inclusion of independent variables in the multivariate model was done if they prove association with the desired outcome, or if their removal affected the relationship. All variables found to be associated with the outcome at the p≤ 0.10 level in the univariate analysis were considered for inclusion in the multivariate model. In this study, P-value of less or equal to 0.05 (p≤0.05) was set as the level of statistical significance for the tests performed.

### Ethical considerations

Ethical approval for this study was obtained from the Amhara National Regional State Health Bureau (ANR/H/5.311/47). Our data collection was extraction of health facility based follow-up data, hence, formal consent for this activity was not taken as data wasn’t linked to individuals. Nonetheless, we informed the health facility officials of the research and received oral consent for data extraction.

## Results

### 4.1 Participant characteristics

The study selected 800 women who used ANC follow-up, delivery, postnatal care follow-up, Penta 3 immunizations both from baseline and intervention groups. Equal number of women from the intervention and baseline were included in the study. In this case, 800 women (400 from before and 400 after mHealth initiated) who used maternal and new-born health services 24 months before and after mHealth started in four Health Centers in Habru District were included in the study. The number of study participants randomly selected from each Health Center were determined based on the client flow (Table 2).

**Table 2:** Number of Study Participants by health facility

| Health Center | Before mHealth (%) | After mHealth (%) |
| --- | --- | --- |
| Mersa | 144(50) | 144(50) |
| Wurgesa | 99(50.3) | 98(49.7) |
| Haro | 87(50) | 87(50) |
| Sirinka | 70(49.6) | 71(50.4) |
| **Total** | **400** | **400** |

All study participants were between the ages of 15-49 years. Age for majority of (77.3%) and 74% of study participants from the baseline and intervention groups, respectively were below the age of 30 years. The mean age for the baseline was 25.73(standard deviation [SD] ±5.22) years and the mean age for the intervention group was 26.16(SD ± 5.66) years. Within groups the heterogeneity of age of study participants used maternal and new-born health services before and after start of mHealth exhibited wider age range, with an age range of 30 years between the youngest and the oldest respondents.

The age of 547 (68.4%) participants from both pre and post mHealth were between 20 and 29 years. A further 195(24.4%) of the participants were age within the range of 30 and 45 years. The proportion of age 40 and above were comparable in both the baseline and intervention groups (Table 3). Age wise, the study participants from both the baseline and the intervention groups were comparable (Table 3). Age variation between pre and post mHealth women was observed between ages 30-34 years. Ninety-six (96%) and 97.5% of the baseline and mHealth intervention study participants, respectively were married; 14(3.5%) study participants from the baseline and 10(2.5%) of the intervention were single, and 2(0.5%) of the baseline were divorced (Table 4). Only 14 (3.5%) and 2 (0.5%) women from the baseline were single and divorced, respectively. Likewise, of women from the intervention group, 10(2.5%) were single. Eight percent of women from baseline and 2.0% from intervention had history of consuming Khat during their pregnancy (Table 3).

**Table 3:** Proportion of study participants by background characteristics

| Category |  | Before mHealth<br>(n=400) |  | After mHealth<br>(n=400) |  |  |  |  |
| --- | --- | --- | --- | --- | --- | --- | --- | --- |
|  |  | % | 95%CI | % | 95%CI | t-test | 95%CI | p-value |
| Age<br>( years) | 15-19 | 8 | 6-11 | 6.5 | 4.5-9.4 | -0.8 | -.05,.02 | P>0.05 |
|  | 20-24 | 33 | 29-38 | 37 | 32.4-41.9 | 5.6 | -.03,.11 | P>0.05 |
|  | 25-29 | 36.3 | 32-41 | 35.8 | 31.2-41 | -.15 | -.07,.06 | P>0.05 |
|  | 30-34 | 13.8 | 11-18 | 8.5 | 6.1-11.7 | 2.4 | .01, .1 | P<0.05 |
|  | 35-39 | 7.5 | 5.3-11.0 | 10 | 7.4-13.4 | 1.3 | -.01,.06 | P>0.05 |
|  | ≥ 40 | 1.5 | 0.7-3.3 | 2.3 | 1.2-4.3 | 0.8 | -.01, .03 | P>0.05 |
| Marital Status | Married | 96 | 94-98 | 97 | 96-99 | 1.2 | -.01,.04 | p>0.05 |
|  | Single | 4 | 2.5-6.5 | 3 | 1.4-4.6 | -1.2 | -.04,.01 | p>0.05 |
| Parity of mother | Primipara | 37 | 33-42 | 37 | 32-41 | -.22 | -.06, .08 | 0.83 |
|  | Multipara | 63 | 59-67 | 64 | 59-68 | .22 | -.08, .06 | 0.83 |
| Gestation weeks | < 37 weeks | 13.5 | 9.8-18.2 | 11.1 | 7.8-15.6 | -.78 | -.05,.02 | p>0.05 |
|  | ≥ 37 weeks | 86.5 | 81.8-90.2 | 88.9 | 84.4-92.2 | 0.5 | -.05,.09 | p>0.05 |
| Birth weight | < 2500 gram | 57.0 | 52-62 | 29.8 | 25.5-34.5 | -8 | -.34,-.20 | P< 0.05 |
|  | ≥ 2500 gram | 43.0 | 38-48 | 70.2 | 65.5-74.5 | 8 | .20,.34 | P< 0.05 |
| Preterm | Yes | 13.5 | 9.8-18.2 | 11.1 | 7.8-15.6 | -.82 | -.08,.03 | p>0.05 |
|  | No | 86.5 | 81.8-90.2 | 88.9 | 84.4-92.2 |  |  |  |
| Substance use<br>(Khat) | Yes | 8.0 | 5.7-11.1 | 2.0 | 0.1-4.0 | -3.9 | -.09,-.03 | P< 0.05 |
|  | No | 92.0 | 88.9-94.3 | 98.0 | 96-99 |  |  |  |

**Table 3.1:**
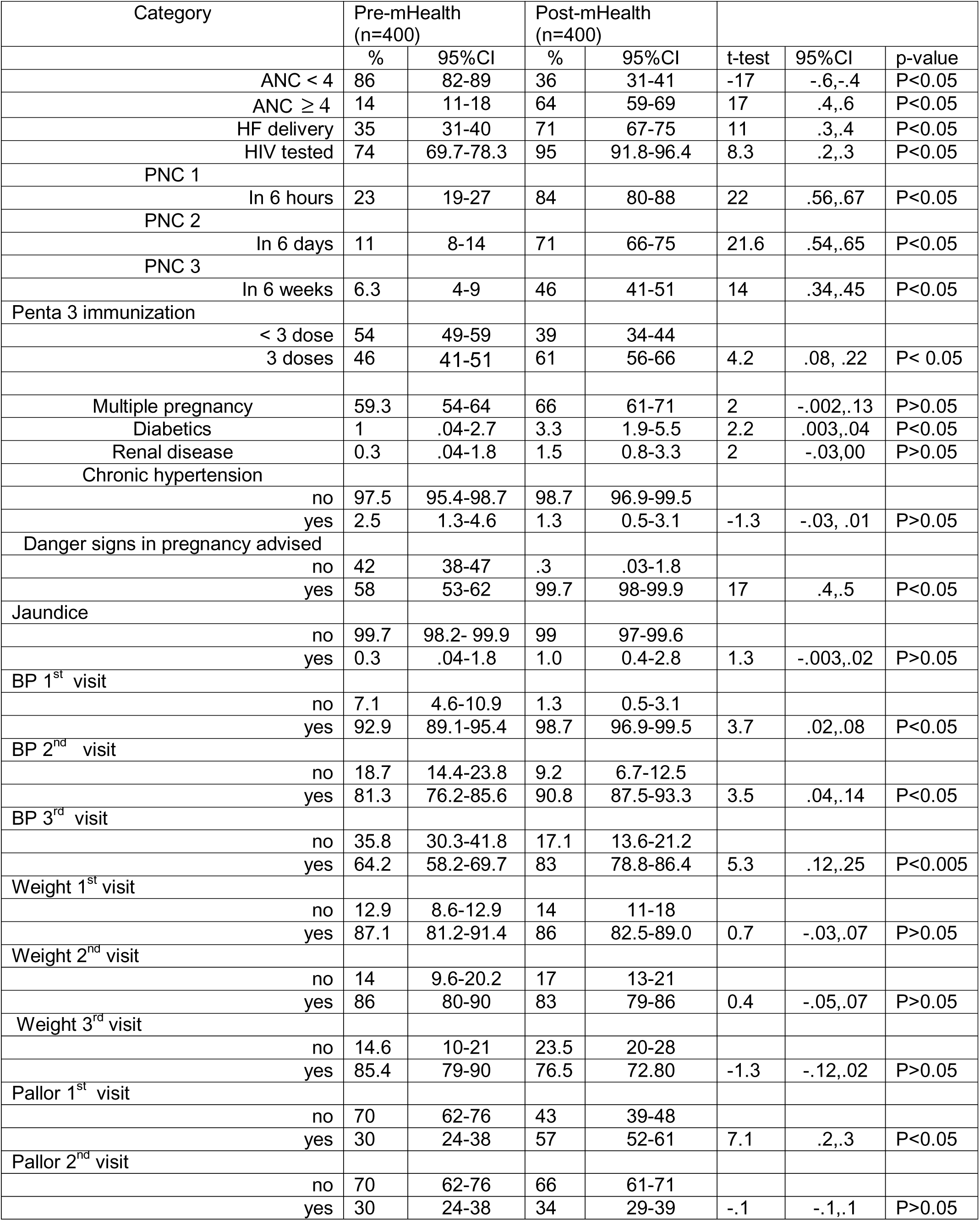
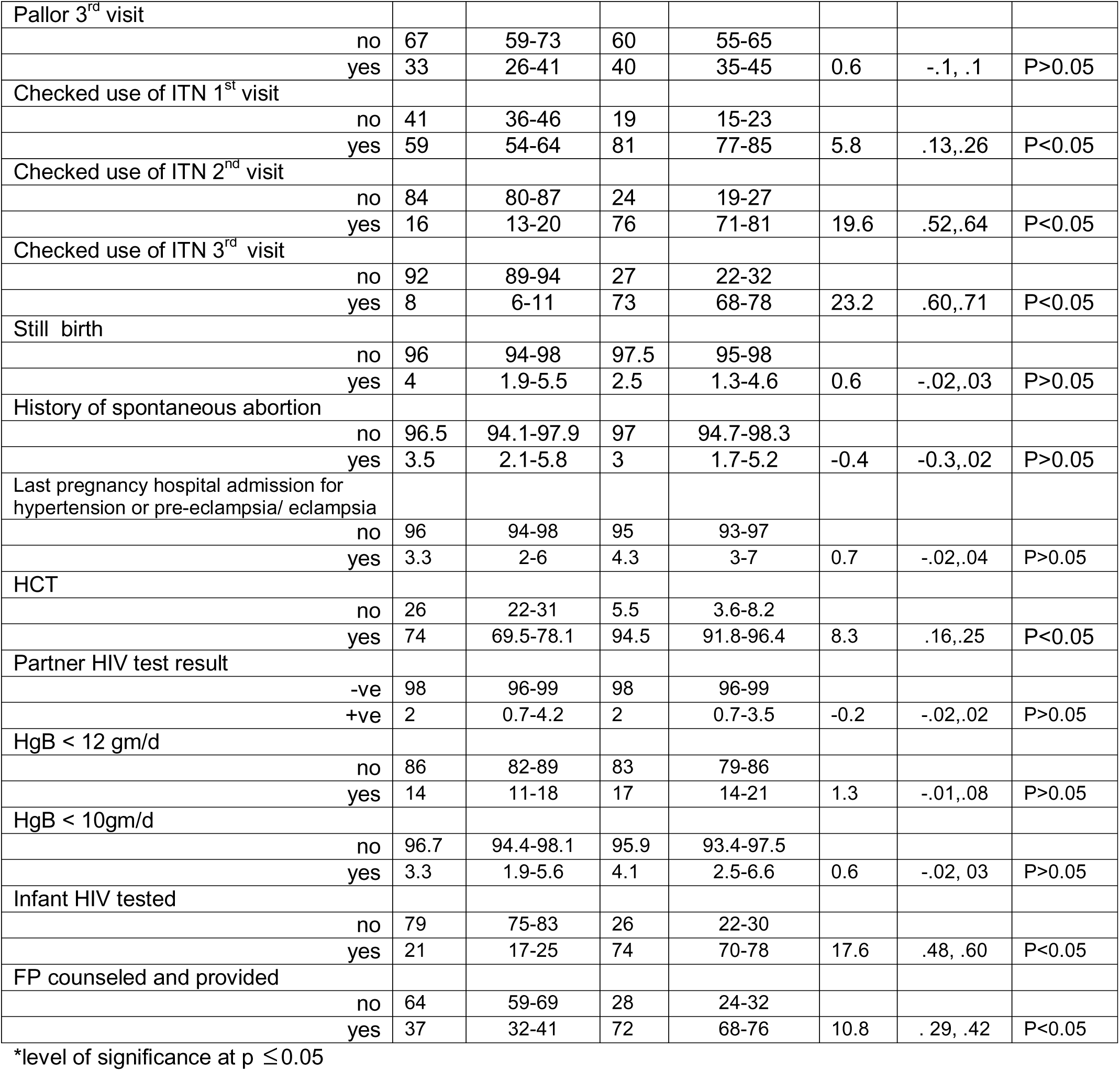
Clinical characteristics and use of maternity services

| Category | Pre-mHealth<br>(n=400) |  | Post-mHealth<br>(n=400) |  |  |  |  |
| --- | --- | --- | --- | --- | --- | --- | --- |
|  | % | 95%CI | % | 95%CI | t-test | 95%CI | p-value |
| ANC < 4 | 86 | 82-89 | 36 | 31-41 | -17 | -.6, -.4 | P<0.05 |
| ANC ≥ 4 | 14 | 11-18 | 64 | 59-69 | 17 | .4, .6 | P<0.05 |
| HF delivery | 35 | 31-40 | 71 | 67-75 | 11 | .3, .4 | P<0.05 |
| HIV tested | 74 | 69.7-78.3 | 95 | 91.8-96.4 | 8.3 | .2, .3 | P<0.05 |
| PNC 1 |  |  |  |  |  |  |  |
| In 6 hours | 23 | 19-27 | 84 | 80-88 | 22 | .56, .67 | P<0.05 |
| PNC 2 |  |  |  |  |  |  |  |
| In 6 days | 11 | 8-14 | 71 | 66-75 | 21.6 | .54, .65 | P<0.05 |
| PNC 3 |  |  |  |  |  |  |  |
| In 6 weeks | 6.3 | 4-9 | 46 | 41-51 | 14 | .34, .45 | P<0.05 |
| Penta 3 immunization |  |  |  |  |  |  |  |
| < 3 dose | 54 | 49-59 | 39 | 34-44 |  |  |  |
| 3 doses | 46 | 41-51 | 61 | 56-66 | 4.2 | .08, .22 | P< 0.05 |
| Multiple pregnancy | 59.3 | 54-64 | 66 | 61-71 | 2 | -.002, .13 | P>0.05 |
| Diabetics | 1 | .04-2.7 | 3.3 | 1.9-5.5 | 2.2 | .003, .04 | P<0.05 |
| Renal disease | 0.3 | .04-1.8 | 1.5 | 0.8-3.3 | 2 | -.03, .00 | P>0.05 |
| Chronic hypertension |  |  |  |  |  |  |  |
| no | 97.5 | 95.4-98.7 | 98.7 | 96.9-99.5 |  |  |  |
| yes | 2.5 | 1.3-4.6 | 1.3 | 0.5-3.1 | -1.3 | -.03, .01 | P>0.05 |
| Danger signs in pregnancy advised |  |  |  |  |  |  |  |
| no | 42 | 38-47 | .3 | .03-1.8 |  |  |  |
| yes | 58 | 53-62 | 99.7 | 98-99.9 | 17 | .4, .5 | P<0.05 |
| Jaundice |  |  |  |  |  |  |  |
| no | 99.7 | 98.2- 99.9 | 99 | 97-99.6 |  |  |  |
| yes | 0.3 | .04-1.8 | 1.0 | 0.4-2.8 | 1.3 | -.003, .02 | P>0.05 |
| BP 1<sup>st</sup> visit |  |  |  |  |  |  |  |
| no | 7.1 | 4.6-10.9 | 1.3 | 0.5-3.1 |  |  |  |
| yes | 92.9 | 89.1-95.4 | 98.7 | 96.9-99.5 | 3.7 | .02, .08 | P<0.05 |
| BP 2<sup>nd</sup> visit |  |  |  |  |  |  |  |
| no | 18.7 | 14.4-23.8 | 9.2 | 6.7-12.5 |  |  |  |
| yes | 81.3 | 76.2-85.6 | 90.8 | 87.5-93.3 | 3.5 | .04, .14 | P<0.05 |
| BP 3<sup>rd</sup> visit |  |  |  |  |  |  |  |
| no | 35.8 | 30.3-41.8 | 17.1 | 13.6-21.2 |  |  |  |
| yes | 64.2 | 58.2-69.7 | 83 | 78.8-86.4 | 5.3 | .12, .25 | P<0.005 |
| Weight 1<sup>st</sup> visit |  |  |  |  |  |  |  |
| no | 12.9 | 8.6-12.9 | 14 | 11-18 |  |  |  |
| yes | 87.1 | 81.2-91.4 | 86 | 82.5-89.0 | 0.7 | -.03, .07 | P>0.05 |
| Weight 2<sup>nd</sup> visit |  |  |  |  |  |  |  |
| no | 14 | 9.6-20.2 | 17 | 13-21 |  |  |  |
| yes | 86 | 80-90 | 83 | 79-86 | 0.4 | -.05, .07 | P>0.05 |
| Weight 3<sup>rd</sup> visit |  |  |  |  |  |  |  |
| no | 14.6 | 10-21 | 23.5 | 20-28 |  |  |  |
| yes | 85.4 | 79-90 | 76.5 | 72.80 | -1.3 | -.12, .02 | P>0.05 |
| Pallor 1<sup>st</sup> visit |  |  |  |  |  |  |  |
| no | 70 | 62-76 | 43 | 39-48 |  |  |  |
| yes | 30 | 24-38 | 57 | 52-61 | 7.1 | .2, .3 | P<0.05 |
| Pallor 2<sup>nd</sup> visit |  |  |  |  |  |  |  |
| no | 70 | 62-76 | 66 | 61-71 |  |  |  |
| yes | 30 | 24-38 | 34 | 29-39 | -.1 | -.1, .1 | P>0.05 |
| Pallor 3<sup>rd</sup> visit |  |  |  |  |  |  |  |
| no | 67 | 59-73 | 60 | 55-65 |  |  |  |
| yes | 33 | 26-41 | 40 | 35-45 | 0.6 | -.1, .1 | P>0.05 |
| Checked use of ITN 1<sup>st</sup> visit |  |  |  |  |  |  |  |
| no | 41 | 36-46 | 19 | 15-23 |  |  |  |
| yes | 59 | 54-64 | 81 | 77-85 | 5.8 | .13,.26 | P<0.05 |
| Checked use of ITN 2<sup>nd</sup> visit |  |  |  |  |  |  |  |
| no | 84 | 80-87 | 24 | 19-27 |  |  |  |
| yes | 16 | 13-20 | 76 | 71-81 | 19.6 | .52,.64 | P<0.05 |
| Checked use of ITN 3<sup>rd</sup> visit |  |  |  |  |  |  |  |
| no | 92 | 89-94 | 27 | 22-32 |  |  |  |
| yes | 8 | 6-11 | 73 | 68-78 | 23.2 | .60,.71 | P<0.05 |
| Still birth |  |  |  |  |  |  |  |
| no | 96 | 94-98 | 97.5 | 95-98 |  |  |  |
| yes | 4 | 1.9-5.5 | 2.5 | 1.3-4.6 | 0.6 | -.02,.03 | P>0.05 |
| History of spontaneous abortion |  |  |  |  |  |  |  |
| no | 96.5 | 94.1-97.9 | 97 | 94.7-98.3 |  |  |  |
| yes | 3.5 | 2.1-5.8 | 3 | 1.7-5.2 | -0.4 | -0.3,.02 | P>0.05 |
| Last pregnancy hospital admission for hypertension or pre-eclampsia/ eclampsia |  |  |  |  |  |  |  |
| no | 96 | 94-98 | 95 | 93-97 |  |  |  |
| yes | 3.3 | 2-6 | 4.3 | 3-7 | 0.7 | -.02,.04 | P>0.05 |
| HCT |  |  |  |  |  |  |  |
| no | 26 | 22-31 | 5.5 | 3.6-8.2 |  |  |  |
| yes | 74 | 69.5-78.1 | 94.5 | 91.8-96.4 | 8.3 | .16,.25 | P<0.05 |
| Partner HIV test result |  |  |  |  |  |  |  |
| -ve | 98 | 96-99 | 98 | 96-99 |  |  |  |
| +ve | 2 | 0.7-4.2 | 2 | 0.7-3.5 | -0.2 | -.02,.02 | P>0.05 |
| HgB < 12 gm/d |  |  |  |  |  |  |  |
| no | 86 | 82-89 | 83 | 79-86 |  |  |  |
| yes | 14 | 11-18 | 17 | 14-21 | 1.3 | -.01,.08 | P>0.05 |
| HgB < 10gm/d |  |  |  |  |  |  |  |
| no | 96.7 | 94.4-98.1 | 95.9 | 93.4-97.5 |  |  |  |
| yes | 3.3 | 1.9-5.6 | 4.1 | 2.5-6.6 | 0.6 | -.02, .03 | P>0.05 |
| Infant HIV tested |  |  |  |  |  |  |  |
| no | 79 | 75-83 | 26 | 22-30 |  |  |  |
| yes | 21 | 17-25 | 74 | 70-78 | 17.6 | .48, .60 | P<0.05 |
| FP counseled and provided |  |  |  |  |  |  |  |
| no | 64 | 59-69 | 28 | 24-32 |  |  |  |
| yes | 37 | 32-41 | 72 | 68-76 | 10.8 | .29, .42 | P<0.05 |

\*level of significance at $p \leq 0.05$

**Table 4:** Proportion of study participants by Marital Status

| Category | Marital status |  |  |  |  |  |
| --- | --- | --- | --- | --- | --- | --- |
|  | Married |  | Single |  | Divorced |  |
|  | Frequency | Percent | Frequency | Percent | Frequency | Percent |
| Before mHealth<br>(n=400) | 383 | 95.75 | 15 | 3.75 | 2 | 0.5 |
| After mHealth<br>(n=400) | 389 | 97.25 | 11 | 2.75 | 0 | 0.0 |

More than one-third of participants (37.2%) from baseline and 33.8% of the mHealth participants were primigravida followed by second gravid women of 35% and 32% from baseline and intervention group, respectively. Maternal clinical background and the magnitude of maternal and newborn health service utilization for both groups is described in Table 3.1.

### 4.2 Obstetric History

Data were enquired about marital status, birth-order, frequency of antenatal check-up, gestational age, complication during pregnancy or childbirth, and other obstetric histories. The outcome indicators in the continuum of care for maternal and new-born health pathway were constructed using number and timing of ANC follow-up, facility based delivery, postnatal care, and penta-3 immunization. In measuring ANC follow-up, number of ANC follow-up, received ANC 4 or more attendances, time each ANC follow-up made, and number of essential ANC services provided were captured. Measurement of postnatal check-up includes timing and frequency following delivery. The child’s immunization construct measure whether the child received penta 3 immunization and number of penta 3 vaccines the child received.

Gestational age was calculated from date of childbirth and last menstrual period of the mother. At enrolment median gestational age of 40(IQR, 39-40) and 39(IQR, 39-40) weeks were recorded for baseline and intervention groups, respectively. The mean gestational age for all study participants was 38.8(SD±4.7) weeks.

Apart from demographic information, data were collected on previous fertility. For three (0.4%) of the study participants age between 15 to 19 years the maternal health service utilization were for their first pregnancy. The majority, 99.6% were multi-gravid and the median gravidity for the merged data was 2(IQR, 1-3). Most women (94.8% of the baseline and 96.8% of the intervention; n=766) were multiparous.

In their previous pregnancies 8(1%) women gave birth under 20 weeks of gestation. Women from both the baseline and intervention groups have equal parity (median, 2; IQR, 1-3). The median number of alive children women from both baseline and intervention have was 2(interquartile range [IQR], 1-3). Two hundred thirty nine (59.8%) of the baseline and 265(66.3%) of the intervention women had experienced multiple pregnancies. At prenatal enrolment 13(3.3%) women from baseline and 17(4.3%) from the intervention had pre-eclampsia; of which, five of the women from the baseline and nine out of seventeen of the intervention were multiparous. The age of 9(69%) of pre-eclamsia cases from baseline were between 20-29 years and 12(71%) cases from end line were age 20-34 years.

In this study, 17 (2.1%) pregnancy associated diabetic cases were recorded that constituted 4(1.0%) from baseline and 13(3.3%) from the mHealth intervention group. The frequency of anaemia including mild, moderate, and severe at baseline and intervention groups were 14% and 17.3%, respectively and below 12gm/dl were prevalent among pregnant women age group 20-29 years.

The prevalence of stillbirths from this study was 30/1000 births or 3% (n = 23). Stillbirth in women served by Health Center implementing mHealth (2.5%; n = 10) was lesser compared to baseline in same Health Center (3.3%; n=13). But the observed difference was non-significant (p = 0.269). The stillbirth rate in both the baseline and intervention groups were higher (5.8%; n =13) and (3.5%; n=9), respectively in grand multiparous women (≥4) than in those with a parity of 1 – 3 (*P*<0.005). Risk factors linked with stillbirth are indicated in Table 5.

**Table 5:** Risk factors associated with stillbirth

| Prenatal risk factors |  | Baseline (N=400) |  | Intervention (N=400) |  |
| --- | --- | --- | --- | --- | --- |
|  |  | Number | % | Number | % |
| Chronic hypertension |  | 10 | 76.9 | 5 | 50.0 |
| Anaemia |  | 2 | 15.4 | 3 | 30.0 |
| Multiple pregnancy |  | 11 | 84.6 | 7 | 70.0 |
| Diabetes mellitus |  | 3 | 23.1 | 5 | 50 |
| Birth weight < 2500gm |  | 11 | 84.6 | 5 | 50.0 |
| Preterm |  | 11 | 91.7 | 6 | 85.7 |
| ANC follow-up | ≤ 3 | 12 | 92.3 | 7 | 87.5 |
|  | ≥ 4 | 1 | 7.7 | 1 | 12.5 |
| UTI |  | 9 | 2.3 | 20 | 5 |
| Institutional delivery |  | 3 | 23.1 | 8 | 100.00 |
| Primigravid |  | 2 | 15.4 | 2 | 25.0 |
| Pre-eclampsia |  | 13 | 3.3 | 17 | 4.3 |

At baseline the major risk factors of stillbirth was chronic hypertension, 10(76.9%), pre-eclampsia, 4(30.8%), diabetes mellitus, 3(23.1%), and anaemia, 2(15.4%) at the time of the last pregnancy. In the intervention group, risk factors reported in stillbirth cases include pre-eclampsia 4 (40%), blood pressure 90mm Hg or more at entry to ANC follow-up in 3(37.5%), diabetes mellitus 5(50.0%), renal disease 5 (62.5%), chronic hypertension 5 (50%), and anaemia in pregnancy in 3 (30.0%) women. Both at baseline and intervention, 84.6% and 87.5% of stillbirths, respectively had history of ANC attendance not more than twice. Table 5 shows the risk factors associated with stillbirths.

Recorded gestational age for 13.5% and 11.1% stillbirths from the baseline and intervention groups, respectively were below 37 weeks. Of 13 women who had history of stillbirth or neonatal death 11(84.6%) gave birth below 2500 gram (Table 5). In this study failure to attend ANC 4 or more were significantly associated with stillbirth or neonatal death (Relative Risk [RR], 1.5, 95% Confidence Interval [CI]: 1.29-1.74) when using merged data.

Pre-eclampsia was recorded in 13(3.3%) of the baseline women and 17(4.3%) of the intervention. The factors associated with pre-eclampsia is depicted in Table 6. At baseline 69.2% and 58.8% of the intervention pregnant women were pre-eclampsia of second gravida

**Table 6:** distribution of pre-eclampsia by some background/clinical characteristics

| Category |  | Baseline |  | Intervention |  |
| --- | --- | --- | --- | --- | --- |
|  |  | Frequency | % | Frequency | % |
| Age (years) | ≤ 35 | 2 | 15.4 | 5 | 29.4 |
|  | ≥ 35 | 11 | 84.6 | 12 | 70.6 |
| Previous stillbirth or neonatal death | Yes | 4 | 30.8 | 4 | 23.5 |
| Parity | Primiparous | 3 | 23.1 | 5 | 29.4 |
|  | Multiparous | 10 | 76.9 | 12 | 70.6 |
| Anaemia | Yes | 1 | 7.7 | 3 | 17.6 |
| Diabetes mellitus | Yes | 3 | 23.1 | 10 | 58.8 |

At enrolment to prenatal care 13(3.3%) pregnant women from baseline and 16(4.1%) from intervention were anemic. Over 69% of anemic pregnant women from both the baseline and intervention groups were age between 20-29 years. Among the parity group 9(81.8%) and 11(69%) multiparous pregnant women from baseline and intervention groups, respectively suffered from anaemia.

Total of 29(3.6%) women, 9(2.3%) from baseline and 20(5%) from intervention group diagnosed having UTI. Six (66.7%) UTI cases from baseline were age between 25-29 years of age. On the other hand, 15(75%) women reported having UTI were between 20-29 years. Nine (100%) of the women with UTI cases at baseline had hemoglobin levels below 10 gm/dl (RR, 8.4; 95%CI: 6.4-11.1). While at the intervention group 17(85%) UTI cases had hemoglobin levels below 10 gm/dl (RR, 6.3; 95%CI: 4.6-8.6).

Women who had Gestational Diabetes with previous stillbirth or neonatal loss (RR 7.9; 95%CI: 3.36-11.87), had history of 3 or more spontaneous abortions (RR, 5.8; 95%CI: 3.59-9.36) and substantially more parous (RR, 1.5; 95%CI: 1.13-3.41). Of women included in the study, 10(2.5%) from the baseline and 5(1.3%) from intervention groups had history of chronic hypertension.

In this study 35 (13.3%) women from baseline and 29 (11.1%) from intervention, experienced preterm births (Table 3). Of 35 preterm infants at baseline the birth weight of 23 (65.7%) were below 2500 gram. Likewise, of 29 preterm infants in the intervention group the birth weight of 10(34.5%) new-born were below 2500 grams. The pattern of preterm birth below 37 weeks in the baseline and mHealth intervention groups is depicted in Figure 1.

The average birth weight of child from the baseline and intervention groups were 2489.97(SD±723.86) grams and 2900.38(SD± 804.88) grams, respectively. Birth weight 2500gm and above reached to 70 % at mHealth from 43% pre-mHealth and the observed difference was significant (t=8, p< 0.05).

In this study preeclampsia (AOR, 3.3, 95% CI=1.5-6.8) and hyperglycemia (AOR 3.1; 95%CI 1.6-5.7) were a risk factor for low birth weight in pregnant women from mHealth intervention group. Multivariate analysis of baseline data indicated women with history of stillbirth or neonatal death (OR, 1.26; 95% CI: 1.14-3.89) and pregnancy associated diabetes cases (OR, 3.4; 95%CI: 1.93-7.8) were at risk to develop pre-eclampsia compared to women without history of stillbirth/neonatal death and diabetes mellitus. At the intervention group women with pregnancy associated diabetes mellitus were 4.5 fold more likely to develop pre-eclampsia compared to non-diabetes women (AOR, 4.5; 95%CI:2.46-8.60). Backward stepping multivariate analysis also disclosed that pre-eclampsia were a risk factor for birth weight less than 2500 gm. Women in the mHealth intervention group with pre-eclampsia were 6 fold (AOR, 2.4; 95%CI: 1.8-6.2) more likely to give birth to neonate less than 2500 gm compared to pregnant women with no history of pre-eclampsia. Hence, low birth weight was neonatal complication anticipated when the mother has pre-eclampsia in pregnant women from the intervention group.

History of 3 or more spontaneous abortions were reported in 14(3.5%) and 10(2.5%) women of the baseline and intervention groups, respectively. Of these, 78.6% women from the baseline and 80% from the intervention were age between 35 and 40 years. In the univariate analysis data from intervention group indicated relationship of spontaneous abortion with fewer ANC follow-up. However, the association didn’t last when entered in a multivariate analysis. It was observed that women in the intervention group age 35 and above (AOR: 2.8; 95% CI: 1.82-8.65) and iron deficiency anaemia (AOR: 2.6; 95%CI: 1.29-6.74) were presented with spontaneous abortion. Whereas, at baseline, age in women 35 and above were associated with spontaneous abortion (AOR: 3.89; 95%CI: 1.7 -9.01). At pre-mHealth diabetic mellitus, chronic hypertension, and preeclampsia were associated with low birth weight (Table 7). At the intervention group diabetic mellitus and preeclampsia showed linkage with low birth weight (Table 7).

**Table 7:** Univariate analysis of maternal health and pregnancy history, risk factors for LBW in Habru, North Wollo

| Variable |  | Baseline |  |  |  | Intervention |  |  |  |
| --- | --- | --- | --- | --- | --- | --- | --- | --- | --- |
|  |  | Birth weight (%) |  | OR | 95%CI | Birth weight |  | OR | 95%CI |
|  |  | <2500 | ≥2500 |  |  | <2500 | ≥2500 |  |  |
| Gestational age | < 37 weeks | 23(65.7) | 12(34.3) | 1.15 | 0.88-1.49 | 10(34.5) | 19(65.5) | 1.15 | (0.88-1.49) |
|  | ≥ 37 weeks | 128(57.1) | 96(42.9) |  |  | 72(31.0) | 160(69) |  |  |
| Diabetes Mellitus | Yes | 4(100.0) | 0(0.0) | 1.77 | 1.62-1.93 | 8(61.5) | 5(38.5) | 2.14 | ( 1.35-3.38) |
|  | No | 223(56.8) | 172(43.5) |  |  | 111(28.8) | 275(71.2) |  |  |
| Chronic hypertension | Yes | 8(80.0) | 2(20) | 1.42 | 1.03- 1.96 | 1(20) | 4(80) | 1.14 | (0.73 -1.78) |
|  | No | 219(56.3) | 170(43.7) |  |  | 118(29.9) | 276(70.1) |  |  |
| Anaemia | Yes | 8(61.5) | 5(38.5) | 1.09 | .07-1.68 | 4(25) | 12(75) | 0.83 | (0.35-1.96) |
|  | No | 217(56.7) | 166(43.3) |  |  | 112(30.2) | 259(69.8) |  |  |
| Abortion | Yes | 9(4.0) | 218(96.0) | 1.36 | 0.47-3.99 | 2(1.7) | 117(98.3) | 0.47 | (0.11-2.12) |
|  | No | 5(2.9) | 167(97.1) |  |  | 10(3.6) | 270(96.4) |  |  |
| Pre-eclampsia | Yes | 13(100) | 0(0.0) | 2.5 | 1.78-3.56 | 12(70.6) | 5(29.4) | 5.7 | (2.03-7.85) |
|  | No | 214(55.4) | 172(44.6) |  |  | 107(28) | 275(72) |  |  |
| Parity | Nulliparous | 2(50) | 2(50) | 0.86 | 0.32-2.30 | 1(25) | 3(75) | 0.86 | (0.09-8.12) |
|  | Multiparous | 144(58.3) | 103(41.7) |  |  | 70(28) | 180(72) |  |  |
| Preterm | Yes | 23(65.7) | 12(34.3) | 1.15 | 0.88-1.5 | 10(34.5) | 19(65.5) | 1.11 | (0.65-1.90) |
|  | No | 128(57.1) | 96(42.9) |  |  | 72(31) | 160(69) |  |  |

Compared to women with no history of chronic hypertension women with history of chronic hypertension at baseline were twice at risk to experience spontaneous abortion (AOR 2.5; 95% CI: 1.7-5.5) as shown in Table 8. Multivariate analysis of data from the mHealth intervention group showed women age above 35 years (P=0.002) and anemic (hemoglobin less than 10 mg/dL) (P=0.045) were at risk for spontaneous abortion (Table 8).

**Table 8:** Predictors of spontaneous abortions at baseline

| Predictor variable<br>(n=388) |  | Univariate analysis |  | Multivariate analysis |  |
| --- | --- | --- | --- | --- | --- |
|  |  | 95%CI | P-value | 95%CI | P-value |
| Pre-eclampsia | Yes | (0.31-1.22) | 0.90 | (0.33-1.52) | 0.99 |
|  | No | 1 | - | 1 | - |
| Multiple pregnancy | Yes | (0.40 -2.98) | 0.28 | (0.38-2.98) | 0.28 |
|  | No | 1 | - | 1 | - |
| Chronic hypertension | Yes | (1.7-5.5) | $P<0.05^*$ | (1.4-6.9) | $p<0.05^*$ |
|  | No | 1 | - | 1 | - |
| ANC follow-up | $\geq 4$ | (0.13 -10.36) | 0.91 | (0.13- 4.35) | 0.91 |
| | $\leq 3$ | 1 | - | 1 | - |
| Preterm | Yes | (0.56-2.12) | 0.18 | 0.56-2.12 | 0.18 |
|  | No | 1 | - | 1 | - |
| Anaemia <10gm/dL | Yes | (2.7-8.6) | $P<0.05^*$ | (2.5-9.5) | $p<0.05^*$ |
|  | No | 1 | - | 1 | - |

\* Significant at $p \leq 0.05$

Multivariate analysis using stepwise backward elimination of pooled data was done to predict linkage of hypertension in pregnancy, multiple pregnancy, preterm birth, number of ANC follow-ups, and number of alive children with spontaneous abortion. After adjustment for confounding factors, it was found that spontaneous abortion was influenced by history of preterm birth (AOR, 3.52; 1.35-9.20). Removal of preterm birth from the model showed association of pregnancy with hypertension (AOR, 4.92; 95%CI: 1.58-9.12) and women having more than two children (AOR, 2.7; 95%CI: 1.04-6.90) were at risk for spontaneous abortion. At the intervention group risk factors associated with spontaneous abortion include older age, hemoglobin below 10 gm/dL, and diabetic mellitus (Table 9).

**Table 9:** Predictors of spontaneous abortions at mHealth intervention

| Predictor variable |  | Univariate analysis |  | Multivariate analysis |  |
| --- | --- | --- | --- | --- | --- |
|  |  | 95%CI | P-value | 95%CI | P-value |
| Age (years) | ≥ 35 | (1.5-6.02) | p<0.05* | (1.82-8.65) | p<0.05* |
|  | < 35 | 1 | - | 1 | - |
| Pre-eclampsia | Yes | (0.89-3.85) | 0.061 | (0.16-6.16) | 0.45 |
|  | No | 1 | - | 1 | - |
| Multiple pregnancy | Yes | (0.43 – 1.53) | 0.99 | (0.31-1.49) | 0.99 |
|  | No | 1 | - | 1 | - |
| Chronic hypertension | Yes | (0.39-2.16) | 1.00 | (0.39-2.16) | 1.00 |
|  | No | 1 | - | 1 | - |
| ANC follow-up | ≥ 4 | (0.44-1.96) | 0.99 | (0.44-1.96) | 0.99 |
|  | ≤ 3 | 1 | - | 1 | - |
| Preterm | Yes | (0.54-2.17) | 0.99 | (0.54-2.17) | 0.99 |
|  | No | 1 | - | 1 | - |
| Anaemia <10 gm/dL | Yes | (0.43 – 1.55) | 0.99 | (1.04-4.95) | 0.045* |
|  | No | 1 | - | 1 | - |
| Diabetes mellitus | Yes | 1.9-6.4 | p<0.05* | 1.8-4.7 | p<0.05* |
|  | No |  | - |  |  |

\* Significant at $p \leq 0.05$

In the multivariate analysis of the baseline data, the factors that remained significantly associated with preterm birth were previous history of stillbirth (AOR, 3; 95%CI; 2.66-9.41) as indicated in Table 10. Hypertension (AOR, 2.2; 95%CI: 1.93-4.98) were also a predictor of preterm birth following adjustment by removing women with history of stillbirth from the model.

**Table 10:**
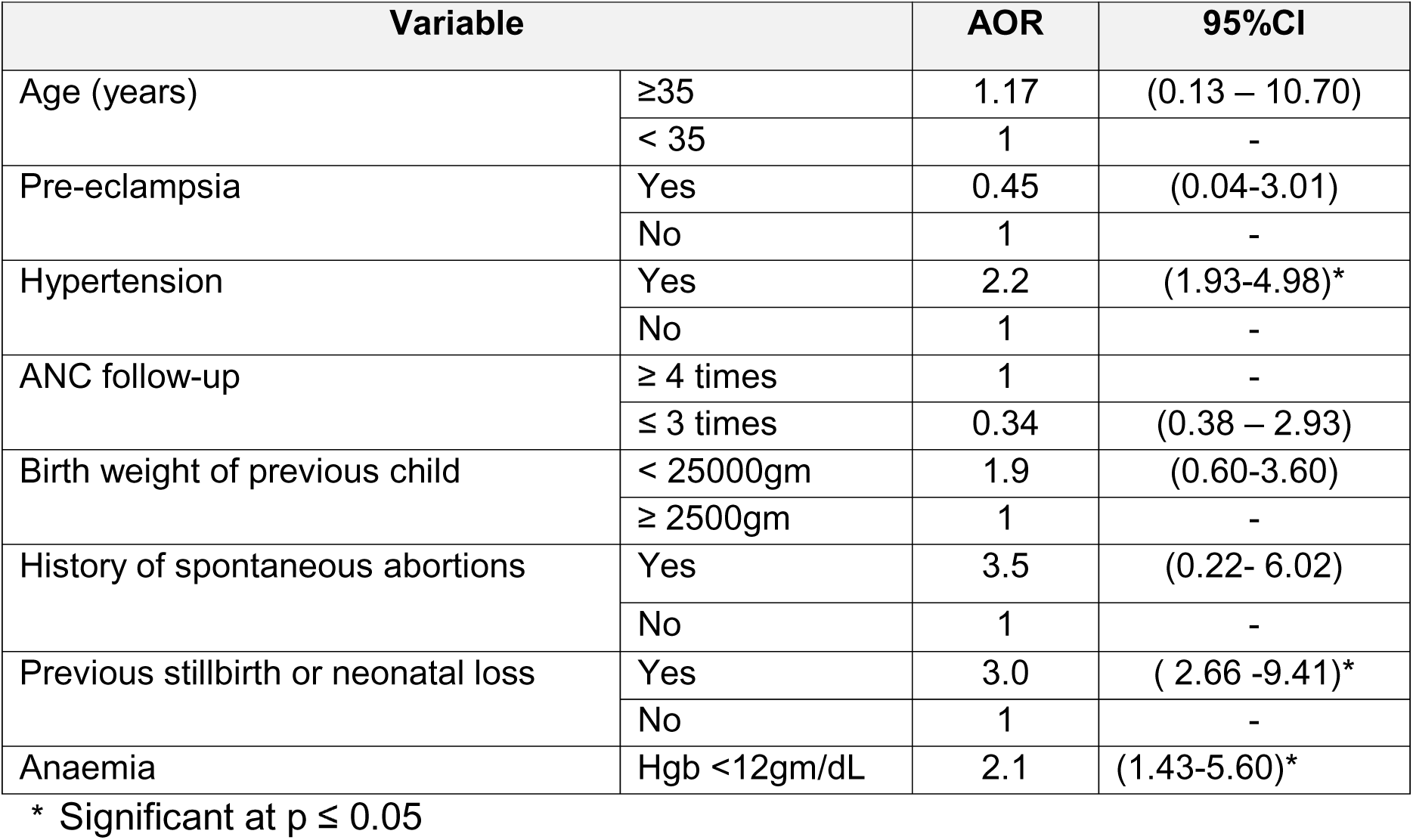
Multivariate analysis on risk factors for early preterm labour at baseline

| Variable |  | AOR | 95%CI |
| --- | --- | --- | --- |
| Age (years) | ≥35 | 1.17 | (0.13 – 10.70) |
|  | < 35 | 1 | - |
| Pre-eclampsia | Yes | 0.45 | (0.04-3.01) |
|  | No | 1 | - |
| Hypertension | Yes | 2.2 | (1.93-4.98)* |
|  | No | 1 | - |
| ANC follow-up | ≥ 4 times | 1 | - |
|  | ≤ 3 times | 0.34 | (0.38 – 2.93) |
| Birth weight of previous child | < 25000gm | 1.9 | (0.60-3.60) |
|  | ≥ 2500gm | 1 | - |
| History of spontaneous abortions | Yes | 3.5 | (0.22- 6.02) |
|  | No | 1 | - |
| Previous stillbirth or neonatal loss | Yes | 3.0 | ( 2.66 -9.41)* |
|  | No | 1 | - |
| Anaemia | Hgb <12gm/dL | 2.1 | (1.43-5.60)* |

\* Significant at $p \leq 0.05$

In women involved in the intervention group diabetic mellitus was a risk factor for preterm labour (Table 11). Compared to non-diabetic women pregnant women of diabetic cases were 3 or more times at risk to have preterm birth. Women diabetic cases were 6 (OR, 3.1; 95%CI: 1.60-7.98) fold more likely to have preterm birth compared to non-diabetic women. In addition, diabetic mellitus was a risk factor for low birth weight in women used mHealth services. Women with history of diabetes mellitus were 3 fold more likely (OR, 3; 95%CI: 1.4-5.7) to give birth to infants less than 2500gm.

**Table 11:** Predictors of preterm birth at intervention group

| Variable |  | AOR | 95%CI |
| --- | --- | --- | --- |
| Age (years) | ≥ 35 | 0.38 | (0.01-11.84) |
|  | < 35 | 1 | - |
| Pre-eclampsia | Yes | 1.29 | (1.14-4.97) |
|  | No | 1 | - |
| ANC follow-up | ≥ 4 | 0.99 | (0.27-3.68) |
|  | ≤ 3 | 1 | - |
| Birth weight of previous child | < 2500 gm | 0.73 | (0.18-2.99) |
|  | ≥ 2500 gm | 1 | - |
| Previous stillbirth or neonatal loss | Yes | 2.86 | (1.14-4.50) |
|  | No | 1 | - |
| UTI | Yes | 3.31 | (1.33-5.85) |
|  | No | 1 | - |
| Number of children | ≥ children | 1.22 | (0.6 - 2.37) |
|  | one child | 1 | - |
| Diabetes mellitus | Yes | 3.1 | (1.60-7.98) |

At the intervention group risk factors associated with preterm birth were urinary tract infections (AOR, 3.31; 95%CI: 1.33-5.85) and previous stillbirth or neonatal loss (AOR, 2.86; 95% CI: 1.14-4.49). Also, at the intervention group, the risk of women with urinary tract infection to have preterm birth increased by 3 times compared to women without urinary tract infection. In addition, women with history of previous stillbirth were more likely to have preterm birth compared to those with no history of stillbirth (Table 11).

### 4.3 Antenatal care service utilization

In this study, the percentage of missing values was low. The highest proportion of missing values was for Expected Date of Delivery (EDD), which was not available in 27 cases (3.4%).

The principal investigator analyzed data of 800 pregnant women (400 baseline and 400 from intervention) in four Health Centers providing maternal and new-born health services. All 800(100%) pregnant women visited an antenatal clinic during their most recent pregnancy. The mean number of antenatal clinic visits at baseline and intervention women were 2.21(SD±1.02) and 3.43(SD±0.88), respectively. This showed more pregnant women made antenatal care visits following the start of mHealth implementation. Moreover, for those who started care early, the median number of visits was higher compared to those who started late.

At mHealth intervention group ANC 1 to ANC 4 follow-ups increased compared to pre-mHealth and the observed change was significant (Table 12). Likewise, ANC ^≥4^ follow-up increased from 14% at pre-mHealth to 64 % at mHealth intervention (Figure 2) and the observed difference was significant (t=17, p<005) (Table 12).

**Table 12:** ANC service utilization before and after start of mHealth

|  | Pre-mHealth |  | Post-mHealth |  | t-test | 95%CI | p-value |
| --- | --- | --- | --- | --- | --- | --- | --- |
|  | % | 95%CI | % | 95%CI |  |  |  |
| ANC 1 | 44.5 | 39.6-49.4 | 77 | 73-81 | 10 | .26,.39 | $P<0.005$ |
| ANC 2 | 50.5 | 45-56 | 85 | 81-89 | 10.3 | .28,.41 | $P<0.005$ |
| ANC 3 | 64.6 | 57-72 | 99 | 97.6-99.9 | 13 | .29,.40 | $P<0.005$ |
| ANC 4 | 54 | 40-67 | 98.6 | 95.6 -99.5 | 11.7 | .37,.52 | $P<0.005$ |
| ANC $\geq 4$ | 14 | 10-17 | 64 | 59-69 | 17 | .4,.6 | $P<0.005$ |

Of 256 pregnant women, who attended 4 and above ANC follow-ups in the intervention group, 80% of them were age between 20-34 years. Among pregnant women who used ANC 4 and above from the intervention group, 137(54%) had 2-4 children and 159 (62.1%) were multiparous.

The study included 4 Health Centers; Haro Health Center recorded higher (21.8%) ANC 4 and above; Sirinka Health Center performed only 7.1% before initiation of mHealth intervention. Following the start of mHealth Haro and Sirinka Health Centers achieved higher, 82.8% and lower, 36.6% ANC 4 and above, respectively (Table 13). In our study settings ANC at least 4 times follow-up increased by at least by four fold at post-mHealth compared to pre-mHealth.

**Table 13:**
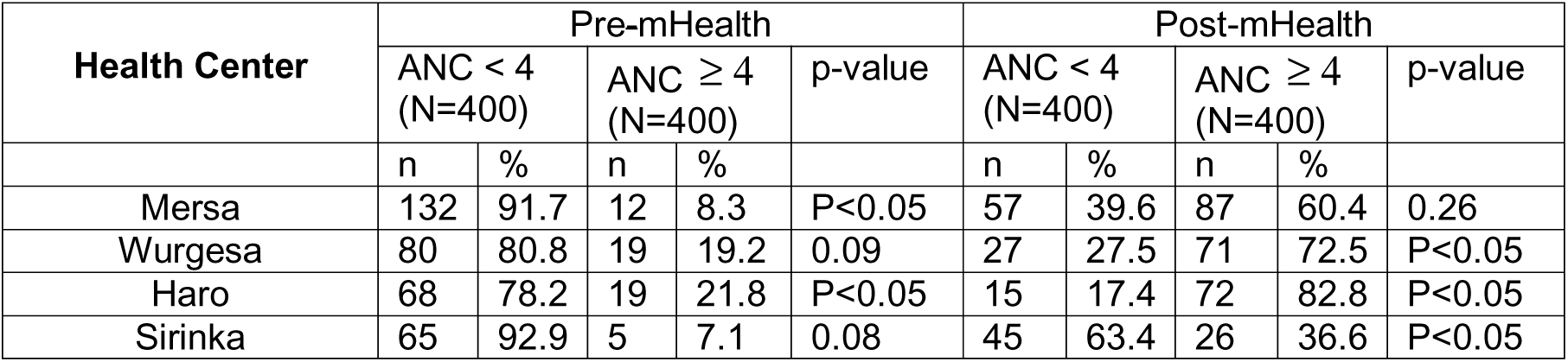
Utilization of ANC across Health centers included in the study

In Health Centers implementing mHealth perinatal care was characterized by a high ANC 4 and above visits, but there are inadequacies that may compromise optimal outcome. Although mean number of follow-up was higher (64%) compared to the baseline (14%) significant proportions of pregnant women (36%) were still classified as having insufficient care. The magnitude of mHealth based ANC 4 and above service uptake varies across Health Centers participated in the study.

At baseline the median time 1^st^, 2^nd^, 3^rd^, and 4^th^ ANC follow-ups made were at 15(IQR, 12-16), 23(IQR, 19-25), 24(IQR, 20-29), and 32(30-34) weeks, respectively. Unlike the baseline the median time in which ANC follow-up started and the number of visits following the initiation of mHealth was at 12, 20, 26, and 30 gestation weeks. Overall, early ANC follow-up at baseline was only 44.5% as compared to the 77.3% of follow ups by pregnant women in the intervention groups. At the intervention group timeliness of prenatal care follow-up increased progressively to 85%, 98.6%, and 99.4% at 2^nd^, 3^rd^, and 4^th^ appointments (table 14 and Figure 3). At mHealth intervention group initiation of ANC 1 and ANC 2 at the recommended time were predictors for use of at least four ANC services. Pregnant women who started ANC 1 at ≥ 12 weeks were 2.5 times (OR, 2.5, 95%CI, 1.5-4.0) and ANC 2 at ≤ 20 weeks of gestation were 7 times (OR, 7.4; 95%CI: 4.4-12.5) more likely to use ANC 4 and above compared to women who started ANC services lately.

**Table 14:** Gestational age at initiation of antenatal care

| ANC visit | Gestational age(weeks) | Before mHealth (N=400 (%)) | After mHealth (N=400) (%) | t-test | 95%CI | p-value* |
| --- | --- | --- | --- | --- | --- | --- |
| 1<sup>st</sup> visit | ≤ 12 | 178(44.5) | 309 (77.3) | 10 | .26, .39 | P< 0.05 |
| 2<sup>nd</sup> visit | ≤ 20 | 144(50.5) | 311(85.2) | 13 | .36,.48 | P< 0.05 |
| 3<sup>rd</sup> visit | ≤ 26 | 106(64.6) | 335(98.6) | 13 | .31,.41 | P< 0.05 |
| 4<sup>th</sup> visit | ≤ 30 | 29(53.7) | 205(99.4) | 11.7 | .37,.52 | P< 0.05 |

\* Independent t-test

The study provides an overview of the services delivered to pregnant women in comparison with the list of services indicated in the ANC chart. As indicated in Table 3.1 service delivery varied in before and after the start of mHealth. Inequalities regarding content of antenatal care were also apparent between the baseline and intervention groups.

While following their antenatal care 296(74%) pregnant women at baseline and 379(95%) at intervention knew their HIV sero-status. All HIV tested pregnant women from the baseline and intervention groups received their test result. Of those HIV tested pregnant women, 5(1.7%) from the baseline and 7(1.8%) from the intervention were HIV sero-positive. All HIV positive pregnant women were counselled on infant feeding and linked to chronic HIV care. Slightly less, 285 (71.3%) partners of pregnant women from the baseline and 377(94.3%) partners from the intervention took the HIV testing and counselling service. Of these, 5(1.8%) from the baseline and 6(1.6%) from the intervention were HIV sero-positive. The HIV test result of one partner of a pregnant woman in the mHealth group was reported as indeterminate. Four out of five HIV seropositive women from the baseline and six-out of - seven from the mHealth intervention group were aged between 20-29 years old.

Additional visits may be necessary for HIV positive women who are on HIV chronic care. Four out of five HIV positive pregnant women from the baseline attended antenatal care only up to three follow-ups. Likewise, six out of seven HIV sero-positive pregnant women from the mHealth intervention group made at least four antenatal visits. All five HIV sero-positive pregnant women from baseline and three out of seven from mHealth intervention were from Mersa Health Center.

Both at baseline and intervention groups antenatal care utilization 4 and beyond were predicator for HIV testing. At baseline pregnant women who used ANC 4 and above were 3 time more likely (OR, 3.1; 95%CI: 1.03-9.1) to opt for HIV testing compared to women who had made fewer ANC follow-ups. Similarly, pregnant women from the intervention groups who attended ANC≥ 4 were 9 fold highly likely to take HIV testing (OR, 9.5; 95%CI: 1.1- 16.4) compared to pregnant women who made fewer ANC visits.

An independent samples t-test was conducted to compare the magnitude of HIV testing in pregnant women at baseline and mHealth intervention groups. There was a significant difference in scores of uptake of HIV testing; baseline (M=0.74, SD=0.45) and mHealth intervention (M=0.95, SD=0.23) conditions. Proportions of pregnant women received HIV testing increased from 74% at pre-mHealth to 95% at post-mHealth and the observed difference was significant (t=8.3; p< 0.05).

#### 4.1.1. Institutional delivery

Baseline and intervention groups had different proportions of unattended home delivery and there were sharp differences between baseline and mHealth intervention group in proportions of facility delivery (35.0% vs. 71.2%).

Of those institutional deliveries 3(3.6%) and 4(1.8%) from baseline and mHealth intervention groups, respectively were stillbirths. In this study, strong linkage was observed between antenatal care and institutional delivery. The frequency of ANC visits was found positively correlated with institutional delivery; the odds of institutional delivery was higher among those who visited ANC at least 4 times (Figure 4).

This study found that institutional delivery at mHealth intervention were higher (0.71 ± 0.45) compared to baseline (0.35±0.49). This difference was significant t (791) = 11; and it did represent a large-sized effect, r = 0.40; p <0.05.

#### 4.1.2. Postnatal care and Penta 3 immunization services

At baseline, of women who gave birth 68(49.6%) delivered women received first postnatal care within 6 hours following delivery, 28(20%) in 6 days, and 16(11.5%) within 6 weeks. And only 22(5.5%) women used their three PNC services within the recommended time while 303(75.8%) women didn’t receive none of the PNC services. In the mHealth intervention group 261(91.6%) women gave birth received first postpartum within 6 hours after delivery, 221(77.5%) in 6 days, and 160(56.1%) made their third postpartum visit within 6 weeks after delivery. At the mHealth 169(42.3%) women timely completed the three commended PNC services; however, 58 (14.5%) women done none of the PNC follow-ups.

A logistic regression framework identified the determinants of postnatal care usage. At baseline use of institutional delivery was predictors of PNC use (AOR 4.5; 95%CI 3.7-8.2). At mHealth, however, use of ANC 4 and above and health facility delivery were predictors of PNC use. After adjusting for covariates using multivariable analysis with multivariable logistic regression, at mHealth it was found that pregnant women who used at least 4 antenatal follow-ups were 3.4 times (AOR, 3.4; 95%C.I:1.8-6.2) more likely to use postnatal care services compared to women who made few antenatal follow-ups. Similarly, women who used skilled delivery were 4.2 times (OR 4.2; 95%CI: 2.3-7.5) more likely to use postnatal care services as compared to women who delivered outside of health facilities.

Generally, postnatal care service utilization across Health Centers included in the study indicated better services utilization in the mHealth intervention compared to the baseline (Figure 5). Postnatal care service utilization at the initial visit were higher but tend to decrease in subsequent visits (Figure 5).

Of women who used no less than 4 antenatal care visits in the mHealth intervention group, PNC follow-up reduced to 57% at third visit from 92% of first PNC follow-up in part due to lack of attention and support for post-partum care. The declining time coincided with the time of termination of support from the donor as the project were financed by a non-Governmental organization.

The type of preventive interventions done during the postnatal period differ across the baseline and mHealth intervention groups. However, the observed difference were significant only for post-partum haemorrhage (PPH) first visit. On average, pregnant women included in the mHealth used more PPH first visit (Mean=2.51, standard error [SE] = 0.043) than the baseline (Mean= 1.25, SE= 0.032). Hence, there was a difference in the level of PPH round first visit follow-up between the baseline and mHealth intervention groups and the difference was significant t (774) = 23.47, p < 0.05; and it did represent a large-sized effect, r = 0.65.

At mHealth intervention Penta 3 vaccine coverage with 3 doses of immunization increased to 61% from 46.3% of the baseline and the observed difference was significant(t=4.2, p<0.05) . At mHealth intervention group 67% of women used minimum of 4 ANC follow-ups had the infants taken penta 3 immunization (UOR, 1.6; 95%CI: 1.20-2.15) compared to 32.7% of the baseline with same ANC uptakes and penta 3 immunization. However, the association of use of ANC 4 and more services with penta 3 service utilization didn’t last when using multivariate backward regression analysis. Likewise, 83.6% (p<0.0001) of women used institutional delivery from the mHealth intervention group had their infants completed penta 3 vaccination compared to 36.8% of the baseline who used same services.

Multivariate analysis of data from mHealth intervention group indicated institutional delivery was predictor for timely immunization of penta 3 immunization. Women used skilled delivery were 4 (OR, 4.3; 95%CI: 2.7-7.1) times more likely to meet penta 3 immunization schedule.

At postnatal the proportions of newborns received full immunizations increased from 36% at baseline to 92.2% at mHealth and the variation was significant (t=11.6, p<0.05). Similarly, the proportions of newborns had vitamin K improved to 94% after the start of mHealth from 79% of the baseline and change was significant at t=3.1 and p< 0.05.

Infant HIV testing improved to 74.2% in the mHealth intervention from 20.3% of pre-mHealth and the observed change was significant (t=17.7, p<0.05). Of these, 4(5.5%) of the baseline and 7(2.4%) of the infants in the mHealth intervention groups were HIV sero-positive. One in four HIV sero positive women and her infant from the baseline and all seven from the mHealth intervention groups were linked to antiretroviral prophylaxis.

More than 72% of the sampled women from the mHealth intervention group and 36.6% of the baseline had adopted a modern method of contraception. Women who had used more antenatal care and skilled delivery service adopted use of modern contraceptives at a higher rate compared to women with a fewer ANC and gave birth beyond health institutions. Overall, women who received family planning counseling increased from 37% at pre-mHealth to 72% post-mHealth and the observed difference was significant(t=10.8, p<0.05). No significant associations were observed between postnatal care service uptake and post-partum family planning practice.

At baseline use of ANC 4 or more follow-up (OR, 2.2; 95%CI: 1.2-4.1) and PNC one visit (OR, 2.2; 95%CI: 1.3-3.8) were predictors of use of family planning services. Likewise, at the intervention use of skilled delivery services (OR, 2.8; 95%CI: 1.7-4.8) and PNC one visit (OR, 5.9; 95%CI: 3.1-11.2) were indicators of use of post-partum family planning.

## Discussion

Gestational diabetic mellitus is defined as carbohydrate intolerance of variable severity with onset or first recognition during pregnancy that does not meet the diagnostic criteria of overt diabetes [6]. Gestational diabetes contribute to perinatal mortality and morbidity [6]. The incidence of gestational diabetes mellitus were 1% and 3.3% for baseline and mHealth intervention groups, respectively. The pooled prevalence of gestational diabetes mellitus developed at 37 and above weeks of gestation was 3.13%. The majority (71%) were age 20-34 years. The pooled prevalence of hyperglycaemia reported in this study is lower than 5.1% published by Domanski et al [7] and 14.9% documented in Brazilian pregnant population [8]. Pooled data indicated diabetes mellitus was implicated with still birth (AOR 5.2; 95%CI 3.1-9.9), chronic hypertension (AOR 3.6; 95%CI 1.8-5.3), spontaneous abortion (AOR 2.6; 95%CI 1.8-4.7), preterm (AOR 3.1; 95%CI 1.6-7.9), and low birth weight (AOR 3.3; 95%CI 1.3-5.4).

Among 400 pregnant women, at baseline the prevalence of chronic hypertension and hypertensive disorders in pregnancy (preeclampsia /eclampsia) were 2.5% and 3.3%, respectively. Likewise, from 400 pregnant women from mHealth intervention group 1.3% were chronic hypertensive and 4.3% were preeclampsia/eclampsia. The prevalence of chronic hypertension observed in this study fall within the range of 1 to 3 per cent reported elsewhere [9, 10, 11]. Among women at mHealth intervention group, the adjusted odds of having a low birth weight baby was more than four times of women without preeclampsia/eclampsia (AOR 4.6; 95%CI 2.4-8.2) and more than two fold(AOR 2.6; 95%CI 1.4-5.2) that for stillbirths. Of ten women of hypertensive cases at baseline 60% were age 35 years and above (AOR 2.4; 95%CI 1.2-5.3). Similarly, five women with hypertension at the mHealth intervention group were age 20-29 years. Conferring to Motghare, Vaz, Pawaskar, and Kulkarni [12] hypertension may cause fetal growth restriction and increase the risk of pre-eclampsia and, thus, the risk of preterm birth, even when treated.

Out of 800 women 29(3.63% [baseline, 2.3% and intervention, 5%]) reported urinary symptoms. The magnitude of UTI reported in our study were much lower compared to 46.5% reported by Haider et al [13]. Urinary tract infection (UTI) is a common clinical problem, which can involve the urethra, bladder, and kidney [14]. However, if untreated, UTI may cause preterm labour [13].

In 2004, the World Health Organization defined stillbirth as the “death of a foetus before the complete expulsion or extraction from its mother at term, weighing at least 1000 gram and occurring after 28 completed weeks of gestation or having at least 35 cm body length” [15]. Perinatal mortality rate in Amhara Regional State was 44/1000 pregnancies [3].

The 3.0% pooled prevalence (baseline, 3.3% and mHealth intervention 2.5%) of stillbirth in our study is higher compared to 0.78% estimate from Ethiopia [16] and lower compared to 4.4% of the Regional stillbirth rate [3]. The stillbirth rate recorded in this study was also lower compared to stillbirth rate between 4.2% and 7.9% documented elsewhere in Ethiopia by Yifru and Asres [17] The observed magnitude of stillbirth may be due to having fewer ANC follow-up visits and not fully utilizing antenatal care services, which should help to detect high-risk women and provide care ensuring a safe delivery. Unsupervised pregnancy is associated with greater frequency of complications, which may be unrecognized and untreated, or inappropriately treated [18]. Of stillbirth cases documented in our study 92.3% and 70% pregnant women at baseline and mHealth intervention groups, respectively used less than four ANC follow-ups. The stillbirth rate is higher in grand multiparous women and this may be due to complications in pregnancy, maternal disease, and complications in labour among women with history of previous stillbirth. The stillbirth rate was also higher in women with multiple pregnancies in 11 (84.6%) of baseline and in 7 (75.0%) of the intervention group. Age wise disparity exists in women with history of stillbirth in the baseline and intervention groups. More than 60% of stillbirth occurred in women age 32±7 years in both baseline and intervention groups.

Birth weight of 2500 gram and above was recorded in 65.6% of women in the intervention group who attended ANC 4 and more and were higher compared to 13.2% of the baseline with identical birth weight and number of ANC follow-up. In present study, the mothers who did visit antenatal clinic less than 4 recommended follow-ups had 1.25 times (OR, 1.25, 95%CI=1.12-1.39) higher odds of having low birth weight baby as compared to those who did recommended visits when merged data were used. Low birth weight of 54.5% were recorded in pregnant women who made three and less ANC follow-ups [19] that is higher compared to 48.7% of pooled data in our study (baseline, 57% and intervention, 28.5%) of low birth weight in pregnant women.

According to WHO [20] abortion is an important cause of morbidity among women of reproductive age, particularly in developing countries. A spontaneous abortion, or miscarriage, is the natural death of a foetus in the womb and is the most common complication of early pregnancy in humans with a rate of 15-20% among pregnant women [21]. Ahmadi et al [18] documented correlation of anaemia with spontaneous abortion. Advanced age as a major risk factor for spontaneous abortion was reported by Correia et al [21]. Positive association of history of spontaneous abortion with anaemia were also reported by Cueto et al [22]. At baseline history of 3 or more spontaneous abortion was associated with birth weight (AOR: 1.4; 95%CI: 1.13-4.19) that is lower compared to findings reported by Makhlouf et al [23] with odds of 2.2.

The findings of this study is analogous to findings from a randomized controlled trial on evaluation of the impact of mobile telephone support on antenatal attendance [24]. The result showed positive association among women in the intervention group and the number of ANC visits (96.4% in intervention group and 92.3% in the control group, P value: 0.002) [24]). Facility based randomized controlled trial using SMS in Zanzibar showed improved ANC utilization (44%) compared to the control (31%) [24].

Antenatal care utilization recorded in this study were higher compared to 44% of four or more antenatal visits in SMS group and 31% of control group, documented by Lee et al [25]. In short message service based antenatal care follow up in Tanzania (Zanzibar) women who received the reminder were more probable to attend four of more antenatal follow-ups (OR 2.39, 95% CI 1.03– 5.55) than those who received routine antenatal care [25]. Improvement of use of ANC 4 and above documented in this mHealth based setting were better compared to 44% of study by Hall et al [26]. SMS based mHealth Intervention of ANC/PMTCT adherence system in Kenya showed a 3 fold increase in ANC visit [27].

At baseline the median time to start 1^st^, 2^nd^, 3^rd^, and 4^th^ ANC follow-ups recorded in this study varied to WHO recommendation of 12, 20, 26, and 30 weeks of gestational ages [28] for 1^st^, 2^nd^, 3^rd^, and 4^th^ ANC visits, correspondingly. At mHealth intervention group, however, the median time for successive ANC follow-ups at 12, 20, 26, and 30 gestation weeks was in concordance to WHO recommendation [28]. Start of ANC at first or early in second trimester of pregnancy as an indicator for uptake of more ANC services during the entire pregnancy recorded in this study is comparable to other study [29]. Early antenatal care follow-up of 77.3% timeliness recorded in this study exceeded improvement from 43.8% to 59% documented in Thailand by Kaewkungwal et al [29] in a study of reminder based before and after mHealth implementation.

Only 55(14%) of baseline and 256(64%) mHealth intervention pregnant women had completed the World Health Organization recommended 4 antenatal visit [30] at the time of their last pregnancy. In Health Centers included in the study ANC service components were in line with the World Health Organization [30] recommendations. The ANC service includes history taking, physical examinations, laboratory examinations and medication administration (Table 3.1) However, differences in the use of ANC services were observed between the baseline and the mHealth intervention groups (Table 3.1). Tables 3.1 presents a comparative analysis of the differences between the two groups who better received the recommended basic ANC service components. The median for utilization of antenatal care service components at baseline and intervention groups were 25.4(IQR, 12.1-47.4) and 91.7(IQR, 67.8 - 98.7), respectively.

Multivariate analysis of mHealth intervention data indicated antenatal care visit at least 4 times were predictor of institutional delivery. Pregnant women who attended 4 and above ANC follow-up were 5 times more likely (OR, 5.1; 95% CI: 2.6-12.8) to use institutional delivery services compared to women who made fewer antenatal follow-ups. This finding is consistent with other studies done in different setting in Ethiopia [31] and in a population based study in Bangladesh by Pervin et al [32], and by Acharya et al in Nepal [33].

Our study revealed that awareness of danger signs at the time of labour and delivery significantly influenced place of delivery. Of women from mHealth intervention group, and gave birth at health facilities, more than 81% were aware of complications while giving birth compared to 44% of the baseline. It might be due to the information received on labour and delivery related complications and advantages of giving birth at health facilities while attending their antenatal appointments.

Antenatal care is recommended to identify pregnancy complications [32]. Antenatal counselling has been described as one of the four main pillars of the Safe Motherhood Initiative because of its effectiveness in reducing maternal and neonatal mortality [34]. Birth preparedness counselling is one of the components of focused antenatal care. Of pregnant women who received birth preparedness planning and counselling during their antenatal follow-up, over 71% from the mHealth intervention group and 34.6% from the baseline used institutional delivery services.

At baseline pregnant women used minimal content of antenatal care components. A higher use of antenatal care service components was associated with higher rates of ANC four times visits observed at the intervention group (Tables 2.1).

In addition, service rendered on seven ANC service components recommended by the national community based new-born care guideline for Focused Antenatal Care (FANC) deeply differs between the baseline and the intervention groups. Review results suggest a consistently high level of antenatal care utilization for the majority of ANC service components following the implementation of mHealth. This may be due to reminding of pregnant women about the next ANC appointment might contribute to the increase of ANC follow-ups; thereby improving uptake of ANC service components as indicated in Table 3.1.

Despite recommendations for HIV testing at point of care for women with an unknown HIV status, HIV testing levels were low among women who had made fewer ANC follow-ups. This is true of all levels of care, but most especially, at the baseline where HIV testing at point of care were only 74%.

Increasing institutional deliveries is crucial for lowering maternal and neonatal mortality. Institutional deliveries have increased from 5% in 2000, 10% in 2011, and 26% in the 2016 [3]. It has been reported that 71.4% deliveries in Amhara Regional State take place at home and majority of them deliver without the assistance of skilled attendants [3]. According to Ethiopian Demographic Health Survey [3] only 27.1% expectant mothers from Amhara Regional State had institutional delivery. It indicates Ethiopia in general and the regional government in particular have a long way to achieve the target of 90% births attended by a skilled provider to meet 2020 target [4]. In this context, an attempt has been made to examine the pattern of institutional deliveries among pregnant mothers in health facilities implementing mHealth based maternal and child health services.

In SMS based before and after study done by Amoakoh-Coleman et al [35] in Rwanda a 20% increase of institutional delivery were documented. In a study in Tanzania (Zanzibar) pregnant women received SMS information on antenatal care were more likely to have skilled attendance at delivery (OR 5.73, 95% CI 1.51–21.81) than those who received routine prenatal care [31]. In SMS based reminder before and after mHealth study done by Ngabo et al [36] 27% increase of institutional delivery was registered.Of pregnant women, used institutional delivery services from the mHealth intervention group, three-quarters of the women were aged 15-29 years.

Institutional delivery was significantly more observed in primiparous mothers (60.4% for baseline and 74.3% for intervention), which is consistent with other studies [37]. The reason could be better understanding of the advantages of maternal health care in young mothers. In addition, the lower parity mother might have less experience at childbirth and may develop fear about the difficulties during labor. Multiparous mothers who had done their previous deliveries at home might also more likely to deliver at home in their recent pregnancy. The finding on parity and Skilled Birth Attendance (SBA) utilization is consistent with similar studies elsewhere [31]. Women with higher parity have experienced childbirth and take childbirth as a natural process; hence, may not seek SBA at delivery if there is no complication [38].

It has been established that ANC is the first step of continuum of services provided during pregnancy. Mothers who attends ANC are likely to attend health facility for childbirth [31]. This study revealed significant positive association between ANC follow-up and skilled birth attendance at delivery. Women who had made at least 4 antenatal care follow-up were more likely to have skilled birth attendance at delivery. Several other studies also found positive association between ANC attendance and facility delivery [38].

Of 256 pregnant women who attended no less than four visits at the intervention group, 207(80.9%) used institutional delivery services, which is higher compared to 55.4% of the baseline with same ANC visits. Over 54 % (78/144) of women received 1-3 times antenatal care follow-up from the intervention group delivered in health facility that exceeds 31.4% of the baseline with same number of prenatal appointments. Overall, institutional delivery increases as the number of ANC follow-up increases (Figure 4).This finding surpasses mHealth based SMS group with skilled delivery attendance of 60% compared to control group of 47% documented by Lee et al [25]. Use of more antenatal care services as predictor of institutional delivery was also recorded in Bangladesh [39] and Rwanda [40].

According to WHO [41] the postpartum period is 42 days after birth. The postpartum period constitute considerable risks for maternal and neonatal health. Lack of health care follow-up during the postpartum may result in mortality or disability, in addition to missed opportunities to promote healthy behaviours. The first hours and days following birth are the most critical for both mother and neonate, even though those in the postnatal period are paid less attention by skilled care providers compared to those in the antenatal and intra-natal periods [41].

The postpartum care visit may be utilized to counsel mothers on infant care and family planning, encourage breastfeeding, identify and treat medical conditions common to the postpartum period, and manage pre-existing or emerging chronic conditions [42].

The 2016 EDHS found that among women age 15-49 years gave birth 2 years before the survey, the proportion of women had first postnatal check in Amhara Region were 0.9% [3]. Close to nine in ten women (87%) did not receive a postnatal check [3]. At least three additional postnatal contacts are recommended for all mothers and new-born, within 6 hours, within 6 days, and 6 weeks after birth [24]. The study explored whether women recently gave birth received any after delivery service namely, postnatal care and family planning within 48 hours of delivery.

At baseline of women who used ANC 4 or more and institutional delivery services only 29% used PNC services. At mHealth intervention group, of women who used identical services 85.5% women used PNC services following delivery. At mHealth intervention group women reported using ANC 4 or more, institutional delivery, and PNC were higher compared to 34% reported in a continuum care [43]. At baseline use of skilled delivery and at the intervention group use of ANC at least 4 times and institutional delivery were predictors of postnatal care services. Consistent with other studies antenatal care was predictor of postnatal care utilization in both intervention and control groups [44, 45, 46]. In SMS based mHealth before and after study in Nigeria clients received text message appointment reminders were 50% less likely to fail to attend their postnatal appointments [24].

The under-utilization of postnatal care may present missed opportunities for family planning discussion as that postnatal care visit would be a good time for such a dialog and family planning method provision [47]. At baseline service intensity of ANC and postnatal care follow-ups were predictors of use of postpartum family planning, which is consistent with other studies [48]. At mHealth intervention group uptakes of PNC [49] and skilled delivery services were indicators of use of postpartum family planning.

Vaccine preventable diseases remain one of the major causes of morbidity, disability and mortality in African Region. Predominantly measles and neonatal tetanus account for most of the 11.4 million deaths documented every year among children under five years of age [50]. According to WHO [51] pentavalent vaccination visit schedules encompass Penat-1 schedule at 6 weeks of baby’s age, Penta-2 schedule at 10 weeks of baby’s age, and Penta-3 schedule at 14 weeks of baby’s age. Because of social issues and insufficient appreciation for immunization, caregivers forget or ignore the importance of immunizations or completing the entire series of vaccines [52].

In SMS based mHealth implementation in Thailand [29] documented increase of median number of immunizations per child to 5 from 3 and Watterson et al [53] recorded increase of childhood immunization from 34.5% to 44.2% in Kenya that is less compared to our finding. In addition, a 95% immunization coverage was recorded compared to 75 % of the non-intervention group implemented in Zimbabwe [54]. In SMS reminder based before and after mHealth implementation evaluation the first, second, and third dose of BCG/HBV/OPV has a 95%, 98%, and 100% rate achievement compared to 60% of total vaccination rates at baseline [50].

## Conclusions

The mHealth project described in this manuscript showed mHealth strategies contributed for the improvement of maternal new-born health service uptakes in rural settings. Use of low cost mobile phones supported with closer follow-up in rural areas with history of lower coverage of maternal and newborn care showed promising results. We found relationship of introduction of mHealth into the health system and improvement of utilization of services like ANC, institutional delivery, postnatal care, and penta-3 immunizations. Among study participants women enrolled in mHealth were more likely to undergo the recommended four and above ANC follow-ups, use institutional delivery, PNC, and penta-3 services compared to women not enrolled in the mHealth. Similarly, enrollment in to the mHealth also improved HIV testing among pregnant women. Web based mHealth system reinforced communication between the midwife at the Health centers and community based health extension workers and contributed for improved maternal and child health services.

## Data Availability

We have prior agreement with the Health Bureau of the Regional government as data is the property of the regional government.

## Acknowledgements

The authors would like to acknowledge the support received from Amhara Regional Health Bureau, North Wollo Health Office, Habru district Health office, and staffs of Health centers included in the study.

## Author Contributions

Conceptualization, protocol development, project administration, analysis, and write up: Aragie Kassa

Review and editing: Mokgadi C. Matlakala

## Notes

### Competing Interest Statement

The authors have declared no competing interest.

### Funding Statement

The author(s) received no specific funding for this work.

### Author Declarations

Ethical approval for this study was obtained from the Amhara National Regional State Health Bureau (ANR 5.311/47). Our data collection was extraction of health facility based follow-up data, hence, formal consent for this activity was not taken as data was not linked to individuals. Nonetheless we informed the health facility officials of the research and received oral consent for data extraction.

